# Functional network organization is locally atypical in children and adolescents with congenital heart disease

**DOI:** 10.1101/2024.04.19.24306106

**Authors:** Joy Roy, William Reynolds, Ashok Panigrahy, Rafael Ceschin

## Abstract

Children and adolescents with congenital heart disease (CHD) frequently experience neurodevelopmental impairments that can impact academic performance, memory, attention, and behavioral function, ultimately affecting overall quality of life. This study aims to investigate the impact of CHD on functional brain network connectivity and cognitive function. Using resting-state fMRI data, we examined several network metrics across various brain regions utilizing weighted networks and binarized networks with both absolute and proportional thresholds. Regression models were fitted to patient neurocognitive exam scores using various metrics obtained from all three methods. Our results unveil significant differences in network connectivity patterns, particularly in temporal, occipital, and subcortical regions, across both weighted and binarized networks. Furthermore, we identified distinct correlations between network metrics and cognitive performance, suggesting potential compensatory mechanisms within specific brain regions.

## 1 Introduction

Congenital Heart Disease (CHD) is a broad term for congenital anomalies affecting the heart’s structure and function, evident from birth (Gonzalez et al., 2021; Sun, Liu, Lu, Zheng, & Zhang, 2015). It stands as one of the most prevalent congenital defects, impacting nearly 1 in 100 infants annually in the U.S.(Sun et al., 2015). Studies have consistently shown that individuals with CHD are more likely to struggle with behavioral challenges, difficulties in school performance, and may encounter obstacles in their careers, such as job-related mobility difficulties (Kamphuis et al., 2002; Warnes et al., 2008). During adolescence, children born with CHD face an increased risk of developing neurodevelopmental impairments (NDI) compared to term-born children, encompassing various deficits affecting academic performance, language skills, memory, attention and social skills, and behavioral function (Nattel et al., 2017; Russell, Chung, Kaltman, & Miller, 2018). Thus, it is imperative to understand the relationship between CHD and neurodevelopment. Specifically, the ability to characterize and predict neurostructural and neurofunctional development in CHD patients could help mitigate potential negative downstream outcomes affecting overall quality of life with better prognosis and treatment planning (Gonzalez et al., 2021).

Functional connectivity (FC) aims to identify linear temporal correlations between blood oxygen level dependent (BOLD) signals between two spatial regions of the brain (Filippi, Spinelli, Cividini, & Agosta, 2019; Mohanty, Sethares, Nair, & Prabhakaran, 2020; Smitha et al., 2017). FC analyses yield networks, with nodes representing brain regions and edges representing correlations between regions. Applying graph theory metrics to these networks provides insights into underlying models of connectivity (Filippi et al., 2019; Mohanty et al., 2020; Smitha et al., 2017). While these metrics are generally subject matter agnostic, they contribute to a deeper understanding of connectivity patterns. Numerous studies have explored FC in relation to neurological, neurodegenerative, and psychiatric diseases, identifying connectivity differences to typically-developing and normative subjects (Filippi et al., 2019; Mohanty et al., 2020; Smitha et al., 2017).

When constructing network matrices, the choice between weighted and binary networks has downstream effects on network metric values and interpretations. Weighted networks directly assign the raw correlation value between nodes to the edges, leading to maximally large, more complex networks. While simplifying network architecture, binarization inevitably sacrifices information by converting continuous values into an on/off representation (Korhonen, Zanin, & Papo, 2021). Binary networks, nevertheless, offer distinct advantages in pruning edges and simplifying complex connectivity patterns by clearing potentially weak or indirect second-order correlations. Absolute thresholding, a method involving the setting of a fixed-valued correlation strength, preserves edges above this threshold while discarding others. In contrast, proportional thresholding retains a fixed percentage of the strongest edges, ensuring consistent density across subject matrices. Each approach presents trade-offs; weighted thresholding considers the impact of weak edges on network topology, known to influence network topology and dynamics (Korhonen et al., 2021; Santarnecchi, Galli, Polizzotto, Rossi, & Rossi, 2014). While weighted networks overcome the need for arbitrary thresholds, they are more susceptible to signal-to-noise ratio (SNR) fluctuations. Absolute thresholding ensures a minimum connection strength among all patients but may lead to varying edge numbers between individuals and groups, potentially causing erroneous distinctions between groups (Korhonen et al., 2021). Proportional thresholding addresses this issue by controlling for density or network cost, though it does not account for connection strength (Achard & Bullmore, 2007; van den Heuvel et al., 2017). Finally, there is no consensus in the literature indicating optimal threshold selection, and ranges or heuristically selected thresholds are often selected.(Garrison, Scheinost, Finn, Shen, & Constable, 2015).

In this study, we investigated FC in children and adolescents with CHD using graph theory metrics. Our goal was to explore differences in brain connectivity between CHD patients and a normative population, providing insights into functional reorganization associated with CHD. Recognizing the challenges in network and threshold selection, our study deliberately employed both weighted and binarized networks and investigated their comparative results.

## 2 Methods

### 2.1 Participants

Participants were recruited from a single institution. Our exclusion criteria included comorbid genetic disorders, contraindications for MRI (e.g., a pacemaker), and non-English speakers. For healthy controls, study exclusion criteria also included preterm birth and neurological abnormalities (e.g., brain malformations, history of stroke, hydrocephalus). Patients with CHD included a heterogenous mix of cardiac lesions, including hypoplastic left heart syndrome (HLHS), aortic arch abnormalities, d-transposition of the great aorta (d-TGA), and other malformations requiring surgical correction in the first year of life. 143 patients with CHD and 98 healthy controls were initially screened. 69 patients with CHD and 92 controls successfully underwent MR scanning. For this analysis, we excluded individuals over 25 years old and those with less than 4.5 minutes of usable single-sequence BOLD scan time. Patients were prospectively recruited at our institution with Institutional Review Board (IRB) approval and oversight (University of Pittsburgh Institutional Review Board STUDY20060128: Multimodal Connectome Study approval 23 July 2020 and STUDY1904003 Ciliary Dysfunction, Brain Dysplasia, and Neurodevelopmental Outcome in Congenital approval 6 February 2023). Written informed consent was obtained from all participants for being included in the study. We have published previous analyses of this prospectively recruited cohort (Sahel et al., 2023; Schmithorst et al., 2022; Wallace et al., 2023). Exclusion criteria were applied, patients born preterm, and patients who did not undergo cognitive testing.

### 2.2 Preprocessing

Figure 1 shows our processing workflow. Scans underwent preprocessing through a customized pipeline developed using the FMRIB Software Library (FSL) and Python 3.6 (Jenkinson, Beckmann, Behrens, Woolrich, & Smith, 2012). This approach follows previously published BOLD motion correction and quality-control guidelines (Power et al., 2014). The preprocessing included motion correction, skull stripping, and normalization. Additionally, images underwent a 6mm Full Width at Half Maximum (FWHM) spatial smoothing filter and temporal bandpass filtering (0.009 Hz < f < 0.08 Hz). The subsequent steps included regression of motion correction realignment parameters and their first derivatives. Frames with framewise displacement (FD) > 0.5 mm and derivative of the temporal variance (DVARS) > 5 were censored and scrubbed before similarity matrix calculation, ensuring a minimum of 4.5 minutes of usable data for all patients. The MNI152 template was registered to patient space using both linear and nonlinear registrations, and segmentation utilized a modified version of the Automated Anatomical Labeling atlas (AALv3). To address reliability concerns in registration, thalamic subdivisions with a limited number of voxels were amalgamated into whole thalamic regions, maintaining laterality distinctions. Additionally, regions with fewer than 20 voxels were excluded from further analysis (i.e. left & right locus coeruleus, left and right ventral tegmental area, and dorsal & medial raphe nucleus).

**Figure 1.**
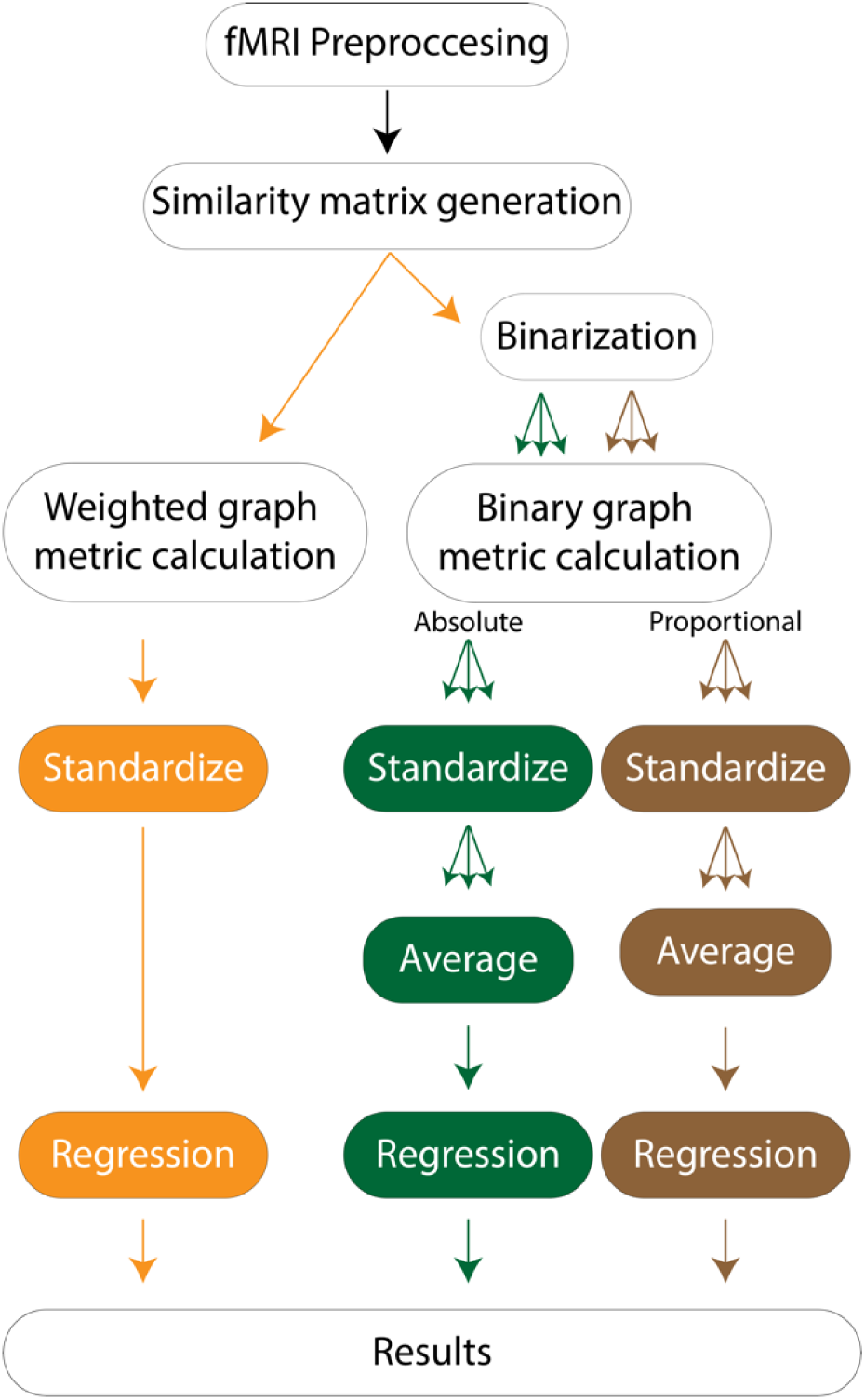
Processing workflow. Each patient’s fMRI scan underwent preprocessing and scrubbing in accordance with the guidelines outlined in Power *et al*. 2014. Similarity matrices were constructed based on the absolute value of the Pearson correlation between regions indicated by the Automated Anatomical Labeling (AAL) template. These matrices were utilized for both weighted analyses and binarization in binary analyses, employing absolute and proportional thresholding iteratively across various thresholds. Subsequently, graph metrics were computed and standardized for each metric and brain region. These metrics served as inputs for regression models predicting neurocognitive outcomes. The metrics were averaged across from all thresholds and were input into regression models. Additionally, we explored regression models at each threshold separately and reported significant results in the Supplementary.

### 2.3 Network Construction

Following preprocessing, the average intensity value for each brain region was computed per timepoint. Pearson correlation coefficients were then calculated between each pair of brain regions over time for each patient, resulting in a patient-specific N x N matrix. Each row and column in this matrix represented a brain region, and each cell held the degree of similarity between the regional temporal BOLD signals. To preserve both strong positive and negative correlations, all matrix values were converted to their absolute values. Subsequently, this square matrix was utilized to construct the adjacency matrix for each patient’s brain network. Our analysis incorporated both a weighted approach and a binary approach, with both absolute and proportional thresholding. The raw matrix served as input for weighted network computations.

To address the arbitrariness of threshold choice in binarization, we adopted an iterative approach, exploring thresholds from 0 to 1 with a step size of 0.05. Constraints were later applied to threshold ranges for both absolute and proportional binarizations to ensure meaningful results. BCTpy’s absolute and proportional thresholding functions were employed for this purpose (Rubinov & Sporns, 2010). Absolute thresholding had a lower bound threshold of 0.10 to mitigate spurious connections and an upper bound threshold of 0.65 to limit the number of components in the network to where the mean and median number of components in the network were less than 2. Proportional thresholding ranged from 0.25, to limit the number of components, to 0.65, where all networks became fully connected.(Supplemental Table 1)

### 2.4 Network Measure Analyses

While numerous software tools are available for network analysis, BCT is specifically tailored for examining network properties in structural and functional connectivity (Rubinov & Sporns, 2010). In this study, we employed BCTpy 0.5.2, a Python implementation of BCT, to compute graph metrics encompassing both weighted and binarized calculations. Nodal efficiency and small-world coefficient were not available in BCTpy and were implemented manually. Across patient networks, a comprehensive set of 14 network measures was computed. Global metrics included global efficiency, assortativity, density, modularity, transitivity, and small-world sigma. Regional metrics were local efficiency, nodal efficiency, clustering coefficient, node betweenness, degree, eigenvector centrality, and participation coefficient. All metrics were calculated per patient network across all thresholds.

#### 2.4.1 Global Network Metrics

Global Efficiency quantifies how efficiently information is exchanged across the entire network. Assortativity indicates the tendency of nodes to connect with others of similar degree. Density reflects the proportion of realized connections to potential connections in the network. Modularity identifies densely connected groups of nodes, revealing community structures. Transitivity measures the degree to which nodes tend to form clusters or triangles. Small World Sigma assesses the balance between segregation and integration in a network.

#### 2.4.2 Regional Network Metrics

Regional metrics include Local Efficiency, gauging how efficiently information is exchanged within the immediate neighborhood of a node, and Nodal Efficiency, identifying nodes crucial for the network’s overall efficiency. Clustering Coefficient related to how connected the network is around a particular node. Node Betweenness quantifies the importance of a node in connecting different parts of the network. Degree quantifies the number of connections each node has. Eigenvector Centrality identifies nodes that connect to other well-connected nodes. Participation Coefficient assesses the diversity of a node’s connections across different network modules.

### 2.5 Executive Function and Network Connectivity

We applied a regression model for each patient to predict their neurocognitive test outcome score, considering factors such as age, sex, presence of CHD, regional network metric, and the interaction between CHD and regional network metric. A total of 7 tests were examined: the National Institute of Health Toolbox (NIHTB) Dimensional Change Card Sort (DCCS), NIHTB Flanker Inhibitory Control and Attention Test, NIHTB Fluid Composite Score, NIHTB Crystalized Composite Score, Wechsler Intelligence Scale for Children IV (WISC-IV) Digit Span test, Behavior Rating Inventory of Executive Function Preschool Version 2 (BRIEF-P) Inhibit subscale, and BRIEF-P Shift subscale. Each test and regional network metric were analyzed separately. Recognizing the diverse range of values for each network metric (e.g., node betweenness ranging from 0 to 1, while node degree spans from 0 to the total number of nodes in the graph), we opted to standardize each metrics across patients before inputting them into the regression model. This standardization process was repeated for both weighted network metrics and for every absolute and proportional thresholded network metric. We then averaged the standardized metric values per region across all thresholds before inputting them into the regression model, as done by Ehrler et al. (Ehrler et al., 2023). To account for multiple comparisons, we focused on results with a coefficient for the interaction term having a p-value < 0.001. In addition, we conducted a separate analysis where, instead of averaging across multiple thresholds, we assessed the performance of each regression model individually on networks trained at each threshold. We report the most significant findings in the Supplementary section. In summary, we calculated regression models from 5 different types of inputs: 1) metrics derived from weighted networks, 2) metrics derived and averaged from several absolute thresholds, and 3) metrics derived and averaged from several proportional thresholds, 4) metrics derived across several absolute thresholds, 5) metrics derived across several proportional thresholds. All analyses used the following regression model:

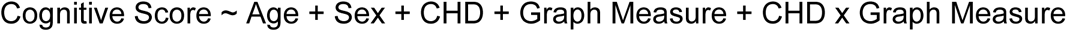

## 3 Results

The dataset initially included 183 patients; however, following the application of exclusion criteria, the total number was reduced to 125 patients. 77 patients and 48 controls had usable BOLD imaging and completed executive function testing. No statistically significant differences in age were observed between CHD and control patients. There was a higher number of control patients compared to CHD patients, and a greater proportion of male CHD patients compared to female CHD patients. Difference in cognitive testing performance between both cohort is listed in Table 1.

**Table 1.** Differences in Cognitive Exam Scores between CHD and Control.

| Outcome | Avg ± Std Control | Avg ± Std CHD | p value |
| --- | --- | --- | --- |
| WISC-IV Digit Span | 10.6 ± 3.0 | 9.2 ± 3.0 | 0.03 |
| NIH Fluid Composite | 106.1 ± 21.7 | 97.9 ± 18.3 | 0.03 |
| NIH Flanker Inhibitory Control and Attention Test | 100.4 ± 13.6 | 96.1 ± 14.4 | 0.10 |
| NIH Dimensional Change Card Sort | 101.3 ± 15.4 | 94.7 ± 16.5 | 0.02 |
| NIH Crystallized Composite | 115.9 ± 15.6 | 105.9 ± 13.4 | 0.00 |
| BRIEF-2 Parent Report Shift | 45.2 ± 8.0 | 49.4 ± 12.1 | 0.05 |
| BRIEF-2 Parent Report Inhibit | 46.2 ± 6.4 | 48.7 ± 7.5 | 0.11 |

Prior to binarization, we saw no significant difference in the average functional connectivity values between CHD and control groups. After adjusting for age and sex, we observed no significant differences in global network connectivity based on metrics such as global efficiency, assortativity, density, transitivity, small-world coefficient, or the number of components across all thresholds and weighted networks. Furthermore, these metrics did not yield a significant coefficient in the interaction term during fitting to the neurocognitive batteries. In both cohorts, networks exhibited assortative behavior and demonstrated small-world characteristics. However, it is noteworthy that the small-world coefficient exhibited a decreasing trend as the network became denser with lighter thresholds (Supplemental Figure 2).

The detected differences in our assessment of neurocognitive outcomes were predominantly in local measures of specific brain regions rather than in global measures across the entire brain network. The outcomes of the regression analyses using weighted metrics are shown in **Table 2**, absolute metrics are presented in **Table 3**, and proportional metrics in **Table 4**. Each coefficient signifies the change in the predicted outcome score for a one-unit alteration. Figure 2 shows regions exhibiting significant interaction terms (i.e., differing relationships between graph measures and cognitive scores in the presence of CHD) and the corresponding cognitive exams. Specifically, eigen vector centrality in temporal brain regions demonstrated variations with the WISC-IV Digit Span across weighted and proportionally thresholded networks. Moreover, eigen vector centrality in the right Lobule IV and V of the cerebellar hemisphere displayed discrepancies with the BRIEF-2 Inhibition score across weighted and proportionally thresholded networks, and the nodal efficiency correlated with the NIHTB DCCS scores in proportional networks. Additionally, the right thalamus exhibited differences with the BRIEF-2 Shift in degree, nodal efficiency, and eigen vector centrality across proportionally and absolute thresholded networks. The Cuneus also demonstrated differences with the BRIEF-2 Shift across degree, nodal efficiency, and eigen vector centrality, with eigen vector centrality showing significant interaction across weighted, proportionally thresholded and absolutely thresholded networks. Overall, these findings reveal patient differences among graph measures in temporal, subcortical, cerebellar, and occipital brain regions across several EF-associated cognitive measures.

**Figure 2.**
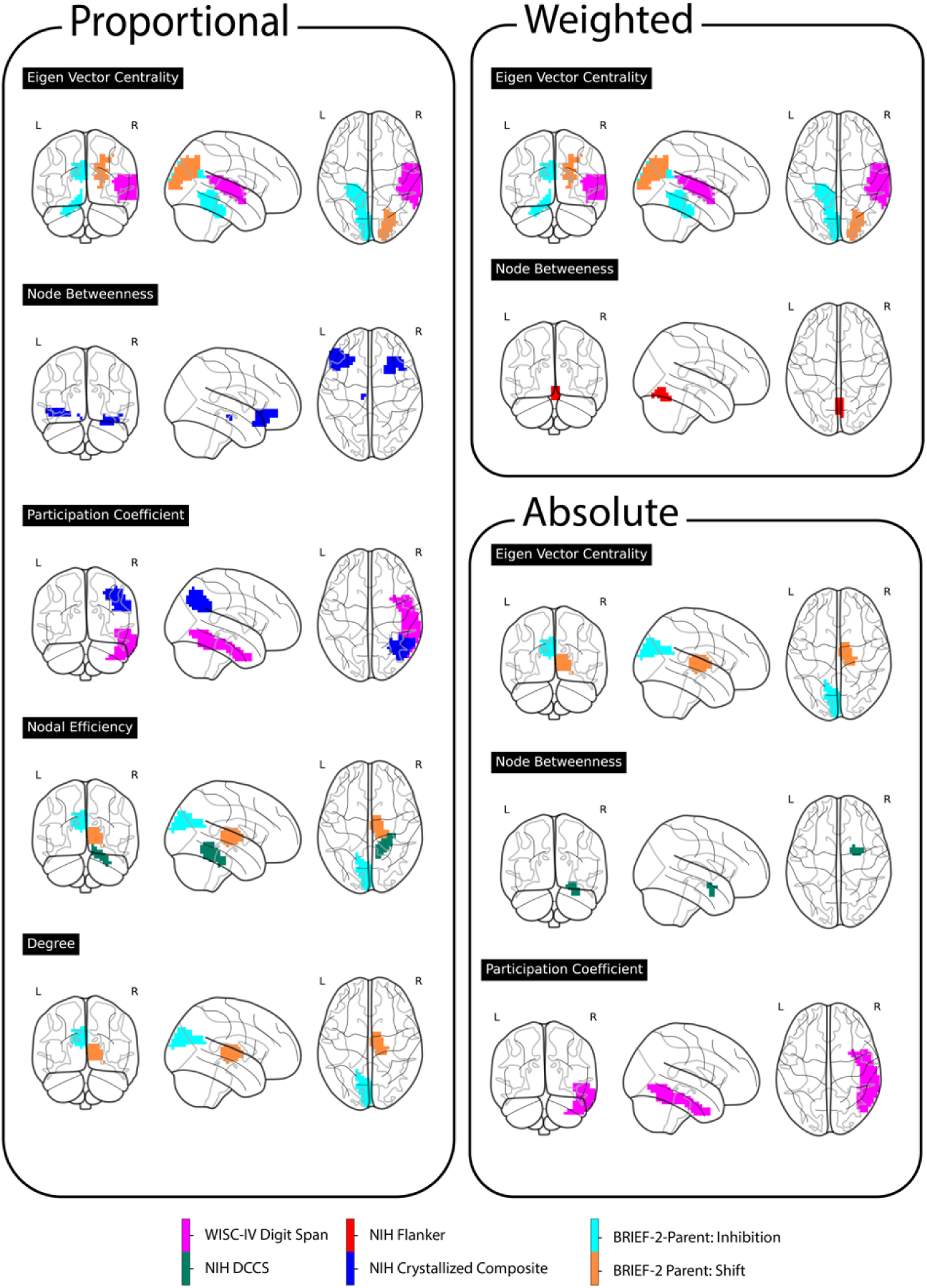
Neurocognitive Exams and Brain Regions. *Figure 3* shows regions with significant interaction effects, indicating varying associations between graph measures and cognitive scores in the presence of CHD, alongside corresponding cognitive assessments. Notably, eigen vector centrality in temporal brain regions showed fluctuations with the WISC-IV Digit Span across two methods. Furthermore, discrepancies in eigen vector centrality in the right Lobule IV and V of the cerebellar hemisphere were observed with the BRIEF-2 Inhibition score across two methods. The right thalamus displayed differences with the BRIEF-2 Shift across three graph measures and two methods. Similarly, the Cuneus exhibited variations with the BRIEF-2 Shift across three graph measures, particularly with eigen vector centrality showing inconsistencies across all three methods. These results underscore patient-specific differences in graph measures across temporal, subcortical, and occipital brain regions across various cognitive assessments.

**Table 2.** Regional Weighted Network Correlations with Neurocognitive Testing.

| Neurocognitive Exam | Brain Region | Network Measure (NM) | NM Coefficient | NM p | CHD Coefficient | CHD p | Interaction Coeff | Interaction p |
| --- | --- | --- | --- | --- | --- | --- | --- | --- |
| WISC-IV Digit Span | Right Superior temporal gyrus | Eigen Vector Centrality | -0.76 | 0.06217 | -1.38 | 0.03433 | 2.26 | 0.00044 |
| NIH Flanker Inhibitory Control and Attention Test | Lobule VI of vermis | Node Betweenness | 2.75 | 0.04040 | -5.10 | 0.04524 | -11.25 | 0.00047 |
| BRIEF-2 Parent Report Shift | Right Superior occipital gyrus | Eigen Vector Centrality | -3.23 | 0.00830 | 5.19 | 0.01501 | 6.90 | 0.00090 |
| BRIEF-2 Parent Report Inhibition | Left Lobule IV, V of cerebellar hemisphere | Eigen Vector Centrality | -2.00 | 0.01348 | 2.79 | 0.05363 | 5.41 | 0.00014 |
| BRIEF-2 Parent Report Inhibition | Left Cuneus | Eigen Vector Centrality | -2.66 | 0.00187 | 3.56 | 0.01492 | 5.85 | 0.00014 |

**Table 3.**
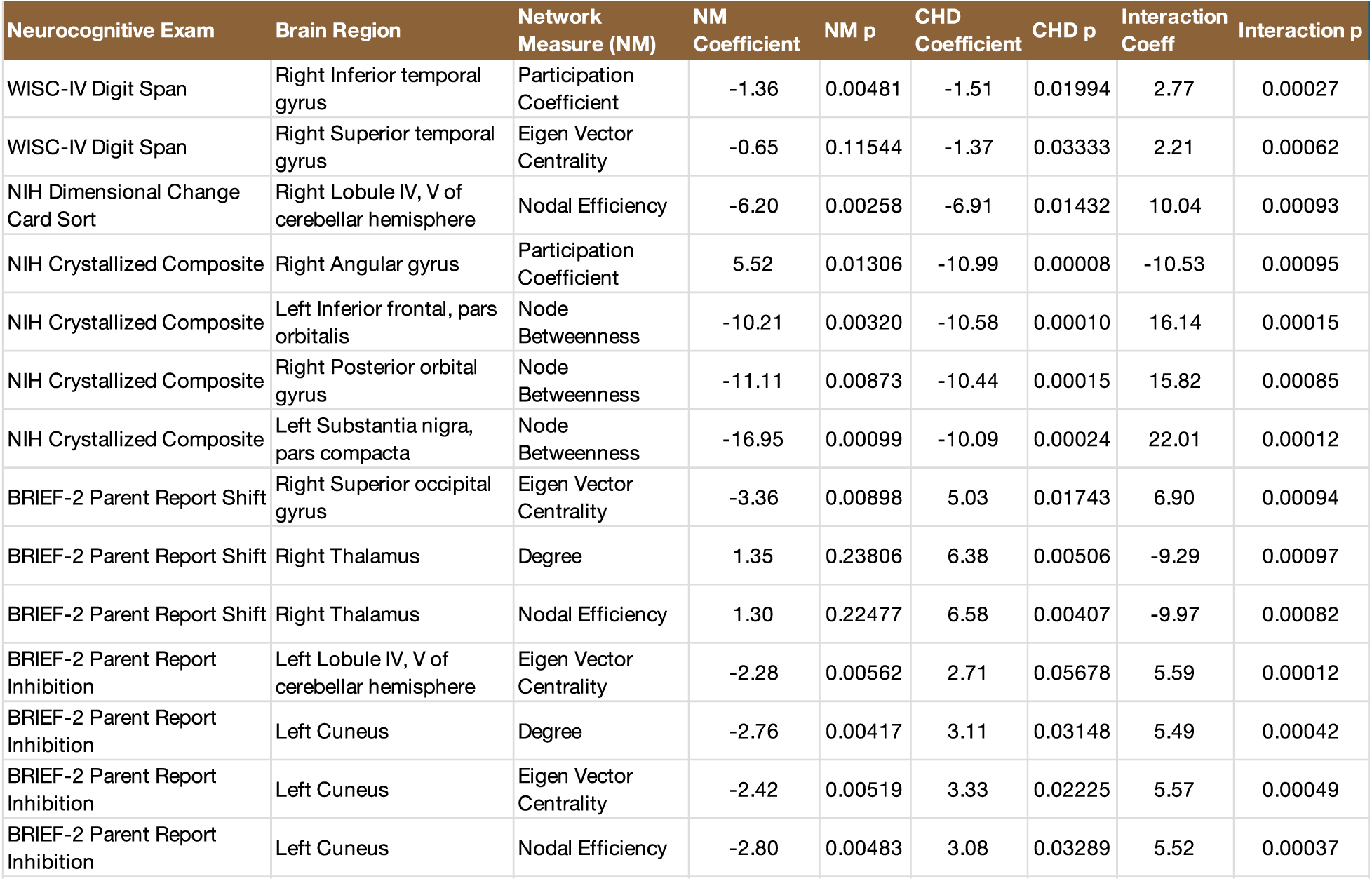
Proportional Thresholding Correlations with Neurocognitive Testing.

| Neurocognitive Exam | Brain Region | Network Measure (NM) | NM Coefficient | NM p | CHD Coefficient | CHD p | Interaction Coeff | Interaction p |
| --- | --- | --- | --- | --- | --- | --- | --- | --- |
| WISC-IV Digit Span | Right Inferior temporal gyrus | Participation Coefficient | -1.36 | 0.00481 | -1.51 | 0.01994 | 2.77 | 0.00027 |
| WISC-IV Digit Span | Right Superior temporal gyrus | Eigen Vector Centrality | -0.65 | 0.11544 | -1.37 | 0.03333 | 2.21 | 0.00062 |
| NIH Dimensional Change Card Sort | Right Lobule IV, V of cerebellar hemisphere | Nodal Efficiency | -6.20 | 0.00258 | -6.91 | 0.01432 | 10.04 | 0.00093 |
| NIH Crystallized Composite | Right Angular gyrus | Participation Coefficient | 5.52 | 0.01306 | -10.99 | 0.00008 | -10.53 | 0.00095 |
| NIH Crystallized Composite | Left Inferior frontal, pars orbitalis | Node Betweenness | -10.21 | 0.00320 | -10.58 | 0.00010 | 16.14 | 0.00015 |
| NIH Crystallized Composite | Right Posterior orbital gyrus | Node Betweenness | -11.11 | 0.00873 | -10.44 | 0.00015 | 15.82 | 0.00085 |
| NIH Crystallized Composite | Left Substantia nigra, pars compacta | Node Betweenness | -16.95 | 0.00099 | -10.09 | 0.00024 | 22.01 | 0.00012 |
| BRIEF-2 Parent Report Shift | Right Superior occipital gyrus | Eigen Vector Centrality | -3.36 | 0.00898 | 5.03 | 0.01743 | 6.90 | 0.00094 |
| BRIEF-2 Parent Report Shift | Right Thalamus | Degree | 1.35 | 0.23806 | 6.38 | 0.00506 | -9.29 | 0.00097 |
| BRIEF-2 Parent Report Shift | Right Thalamus | Nodal Efficiency | 1.30 | 0.22477 | 6.58 | 0.00407 | -9.97 | 0.00082 |
| BRIEF-2 Parent Report Inhibition | Left Lobule IV, V of cerebellar hemisphere | Eigen Vector Centrality | -2.28 | 0.00562 | 2.71 | 0.05678 | 5.59 | 0.00012 |
| BRIEF-2 Parent Report Inhibition | Left Cuneus | Degree | -2.76 | 0.00417 | 3.11 | 0.03148 | 5.49 | 0.00042 |
| BRIEF-2 Parent Report Inhibition | Left Cuneus | Eigen Vector Centrality | -2.42 | 0.00519 | 3.33 | 0.02225 | 5.57 | 0.00049 |
| BRIEF-2 Parent Report Inhibition | Left Cuneus | Nodal Efficiency | -2.80 | 0.00483 | 3.08 | 0.03289 | 5.52 | 0.00037 |

**Table 4.**
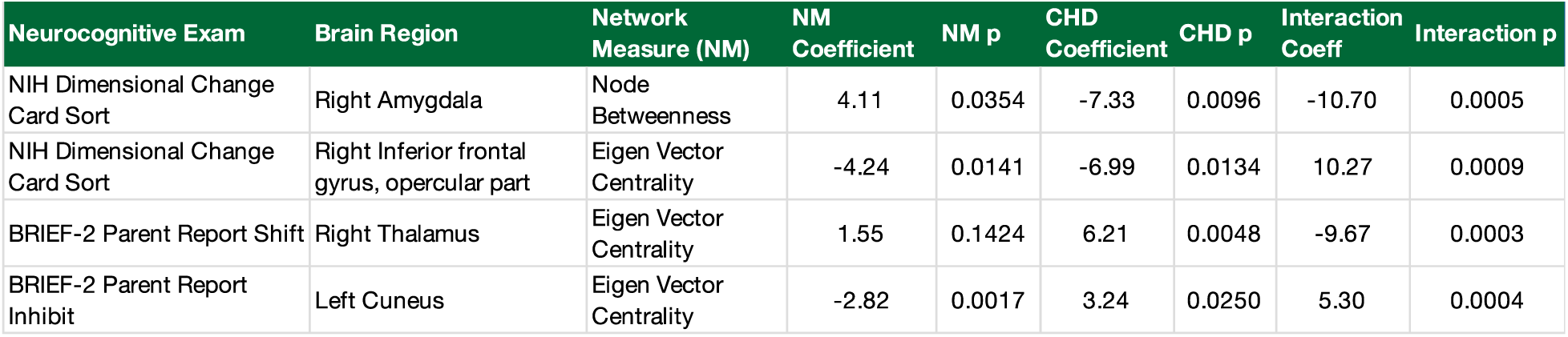
Absolute Thresholding Correlations with Neurocognitive Testing.

Under proportional thresholding, our results suggest that higher nodal efficiency in the Right Lobule IV, V of cerebellar hemisphere is associated with smaller NIHTB DCCS scores. Additionally, the presence of CHD is linked to poorer DCCS scores. Interestingly, the interaction of CHD and higher nodal efficiency in this brain region results in higher DCCS scores. Figure 3 depicts the correlation between each brain region and the predicted effect on cognitive performance with increase in the network metric, considering the presence of CHD (i.e., the interaction term of our regression analyses). Generally, differences are observed in the temporal, occipital, and subcortical brain regions. Regions within the temporal lobe exhibit an overall positive correlation with cognitive performance, whereas the occipital regions show negative correlations. Regarding subcortical regions, the associations are mixed. For instance, the interaction of CHD and node betweenness in the amygdala predicts a decrease in performance. In contrast, the interaction of CHD with degree, nodal efficiency, and eigenvector centrality in the right thalamus predicts an increase in performance.

**Figure 3.**
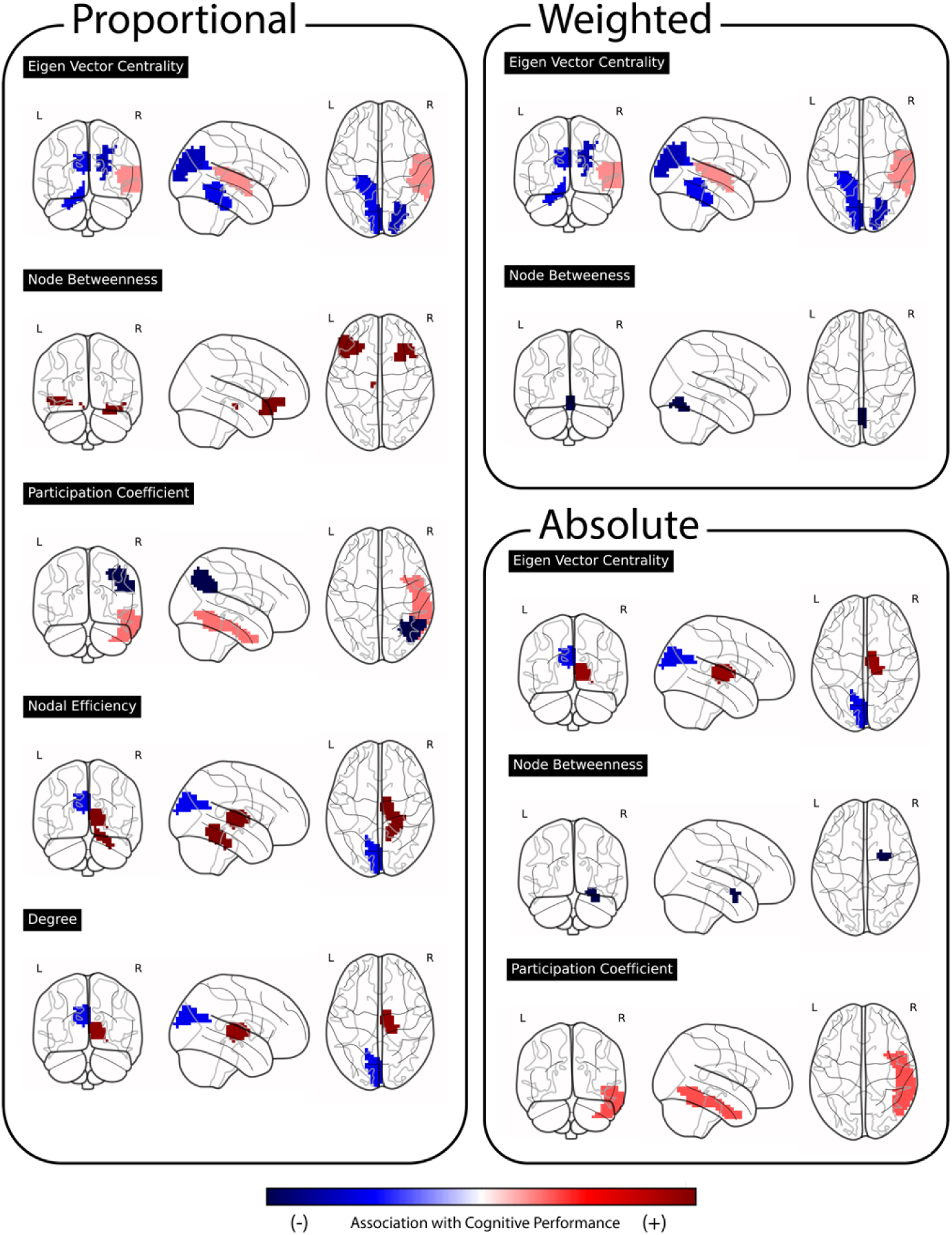
Regional associations with cognitive performance. Temporal lobe regions generally display a positive correlation with cognitive performance, while occipital regions exhibit negative associations. Subcortical regions present a mix of findings, as illustrated by the amygdala’s association with decreased performance when considering the interaction between CHD and node betweenness. Conversely, the right thalamus shows increased performance linked to interactions between CHD and degree, nodal efficiency, and eigen vector centrality.

## 4 Discussion

Here, we used graph metrics to investigate FC differences between CHD survivors and typically developing matched controls. The use of metrics on networks with absolute thresholds emphasizes differences that may be the result of individual functional connection. Proportional metrics, on the other hand, allow for the control of the number of edges while selecting the strongest edges regardless of strength, facilitating an investigation specifically into differences in the topology and distribution of edges. The weighted metrics offer a balanced approach, accounting for both the number and strength of edges, albeit with the acknowledgment that this method may be more susceptible to noise. Our study saw consistent results across methods. For instance, all three methods revealed differences in the interaction of CHD with network metrics in the left cuneus and temporal regions of the brain, indicating variations in both the strength and topology of edges adjacent to these regions. While the cerebellar regions exhibited differences in both weighted and proportional results, they showed no disparity in absolute thresholds. This suggests that while the topology of the cerebellum’s strongest edges may differ, the overall strength of the edges remains consistent. Alternatively, subcortical regions displayed differences in proportional and absolute thresholded networks, but not in weighted networks. This may imply that there are variations in both regional edge strength and topology.

Regression analyses using graph metrics on networks constructed with weighted, absolute, and proportionally thresholded networks highlighted localized discrepancies in neurocognitive outcomes, particularly within specific brain regions, rather than global measures. Notably, differences were observed in the presence of CHD in regions such as the temporal, occipital, and subcortical areas, with varied associations between graph measures and cognitive scores. For instance, interactions between CHD and eigen vector centrality in the right superior occipital gyrus predicted poorer in shift performance in the BRIEF-2 Shift exam, whereas interactions between CHD and eigenvector centrality in the right thalamus predicted improved shift performance, suggesting nuanced impacts on brain function. These results underscore the complex interplay between brain network metrics, cognitive performance, and the presence of CHD.

CHD consistently showed negative correlations with NIHTB and WISC scores, as well as positive correlations with BRIEF scores, suggesting an overall association with decreased cognitive performance. However, the interaction between CHD and certain regional network metrics appears to be linked to improved cognitive performance scores. Specifically, in the temporal regions, the interaction of CHD and a more central area of the right superior temporal gyrus significantly correlates with improved working memory, as assessed by the WISC Digit Span examination. Similarly, a more central and locally efficient right thalamus also demonstrates a positive correlation with better cognitive flexibility seen in BRIEF Shift performance. Together, these findings may suggest potential compensatory or adaptive mechanisms in these brain regions. Conversely, in the occipital regions, such as the right superior occipital gyrus and cuneus areas, alterations in eigen vector centrality were observed to have significant implications for cognitive flexibility and inhibitory control, as assessed by the BRIEF-P Shift and BRIEF-P Inhibit. Decreases in eigen vector centrality within the right superior occipital gyrus and cuneus were associated with diminished cognitive performance in these domains. These findings suggest that more centrally accessible occipital network connectivity may contribute to poorer inhibition and flexibility among CHD patients. Additionally, increases in node betweenness in the amygdala exhibit a large negative correlation with NIHTB DCCS performance, suggesting that the amygdala taking a more central role in connecting other brain regions is associated with poorer executive function. It is worth highlighting in the analysis using absolute thresholded matrices that the right superior temporal gyrus did not meet the p-value cutoff statistical threshold, but notably emerged as the next most significant result, correlating with the WISCV Digit span, displaying an interaction coefficient of 2.17 and a p-value of 0.0011. This finding holds significance as it consistently appeared across all weighted and binarization methods.

We have previously investigated network analyses in CHD patients and their association with cognitive performance, yielding a mix of findings. Panigrahy et al. (2015) utilized diffusion tensor imaging (DTI), graph theory, and statistical mediation models to identify neurocognitive impairments in adolescents who underwent early infancy repair for dextro-transposition of the great arteries (d-TGA), linking these impairments to variations in the overall topology of the white matter structural network (Panigrahy et al., 2015). Similarly, Schmithorst et al. (2016) discovered differences at both global and regional levels, highlighting the role of global white matter structural network topology in mediating adverse ADHD outcomes among adolescents with d-TGA(Schmithorst et al., 2016). Furthermore, Schmithorst et al. (2018) observed variations in regional nodal efficiency, particularly in frontal and subcortical regions, along with global differences in metrics like global efficiency and small-worldness, using structural networks derived from three distinct methods on pre and post operative CHD infants(Schmithorst et al., 2018). These analyses used primarily structural networks whereas our study employed functional networks. Structural connectivity obtained from DTI represents the physical white matter pathways that connect different brain regions. It provides information about the anatomical substrate of brain networks and the pathways through which neural signals propagate. Meanwhile, functional networks reflect the temporal correlations in neural activity between different brain regions. While prior studies indicate structural brain network reorganization, our findings suggest that the global functional network remains largely undifferentiated, with predominantly regional differences. Regional, rather than global differences, may explain higher variance observed in CHD patient scores, but still within normal limits on tests of intelligence (Badaly et al., 2022; Wallace et al., 2023)

Other investigations have similarly identified regional discrepancies in CHD brains while utilizing the NIHTB. The NIHTB has previously found applications in neurocognitive and psychosocial studies within CHD populations (Badaly et al., 2022; Calderon et al., 2019; Cousino et al.; Sahel et al., 2023; Siciliano et al., 2021; Wallace et al., 2023). In a recent comprehensive evaluation of the psychometric properties of the NIHTB-CB and NIHTB-EB among children with CHD, researchers concluded that while the NIHTB may have certain limitations, it remains a valuable tool for CHD analysis and has several correlations and insights (Wallace et al., 2023). A separate investigation revealed a notable connection between changes in regional cerebral blood flow (rCBF) and NIHTB scores among children and adolescents diagnosed with congenital heart disease. Specifically, this study highlighted alterations in the salience network, default mode network, and frontal executive network, indicating potential implications for cognitive functioning in this population (Schmithorst et al., 2022). A separate study used the NIHTB with the same set of patients observed altered cerebello-cerebral connectivity and increased fractional anisotropy correlated with lower scores on the NIHTB tests (Sahel et al., 2023). Another study also delved into the cerebellum of CHD patients and observed associations with cerebellar volume and cognitive test scores (Badaly et al., 2022). The cerebellum was positively associated with greater working memory on the WISC-IV Letter-Number Sequencing; greater working memory on the NIH Toolbox List Sorting Working Memory; mental flexibility on the D-KEFS Trail Making Test; greater inhibitory control on both the D-KEFS Color-Word Interference Test and the NIH Toolbox Flanker Inhibitory Control and Attention (Badaly et al., 2022). Notably, our own research has also identified positive relationship between the cerebellum and working memory in the presence of CHD, seen by a positive correlation between nodal efficiency and the NIHTB Dimensional Change Card Sorting test.

This study has limitations. Functional MRI does not measure brain activity directly, but with the help of BOLD contrast it has become an acceptable and popular way of measuring neuronal activity (Holdsworth & Bammer, 2008). BOLD fMRI relies on blood circulation rather than electrical activity. Thus, disorders affecting blood flow and volume may alter the BOLD signal in healthy regions that otherwise have normal neuronal activity(Holdsworth & Bammer, 2008). These limitations affect fMRI as well as downstream FC interpretations. For example, fluctuating alertness, mental state, and sleep disturbances can all affect fMRI and FC (Buckner, Krienen, & Yeo, 2013). Activation of brain regions in tandem indicates functional connectivity and can be indicative of potential structural connections, albeit not always direct (Thiebaut de Schotten & Forkel, 2022). By solely relying on functional data, our study may not capture the comprehensive understanding that could be achieved by integrating structural imaging modalities. A more holistic examination, incorporating structural information from techniques like diffusion magnetic resonance imaging (dMRI), could have offered a more precise perspective on the interplay between brain structure and function in the context of congenital heart defects. Thus, the study’s findings should be interpreted within the context of this inherent limitation, emphasizing the need for future research endeavors to embrace a multimodal approach for a more comprehensive exploration of the neurobiological underpinnings associated with congenital heart defects.

## 5 Conclusion

Our study investigated the relationship between congenital heart disease (CHD), functional brain network organization, and cognitive performance in children and adolescents. Utilizing regression analyses on graph metrics derived from weighted and binarized networks, we identified localized associations between specific brain regions and neurocognitive outcomes. Notably, interactions between CHD and regional network metrics demonstrated nuanced impacts on cognitive function, highlighting potential compensatory mechanisms within certain brain regions. Our findings underscore the importance of considering both global and regional network characteristics in understanding cognitive deficits associated with CHD. Moreover, our study contributes to the growing body of literature exploring neurocognitive impairments in CHD patients. Further research employing multimodal approaches leveraging both functional and structural imaging may provide deeper insights into the biological underpinnings of CHD-related cognition.

## Data Availability

All data produced in the present study are available upon reasonable request to the authors.
The code used in this study is openly available and can be accessed at the following GitHub repository:

https://github.com/overjoyroy/Sim_Funcky_Pipeline

## Data and Code Availability

The code used in this study is openly available and can be accessed at the following GitHub repository: https://github.com/overjoyroy/Sim_Funcky_Pipeline.

## Author Contributions

JR: Conceptualization, Methodology, Software, Formal analysis, Investigation, Visualization, Writing - Original Draft, Writing - Review & Editing

WR: Methodology, Software

AP: Conceptualization, Writing - Review & Editing, Supervision, Project administration, Funding acquisition

RC: Conceptualization, Methodology, Writing - Review & Editing, Supervision, Project administration, Funding acquisition

## Acknowledgements

We express our sincere appreciation to Julia Wallace for her invaluable support in managing the imaging data. Our heartfelt gratitude extends to the patients and families who generously contributed to this study, as well as to the dedicated PICU staff who deliver exceptional care to post-cardiac arrest patients and their families.

## Supplement

**Supplemental Figure 1.**
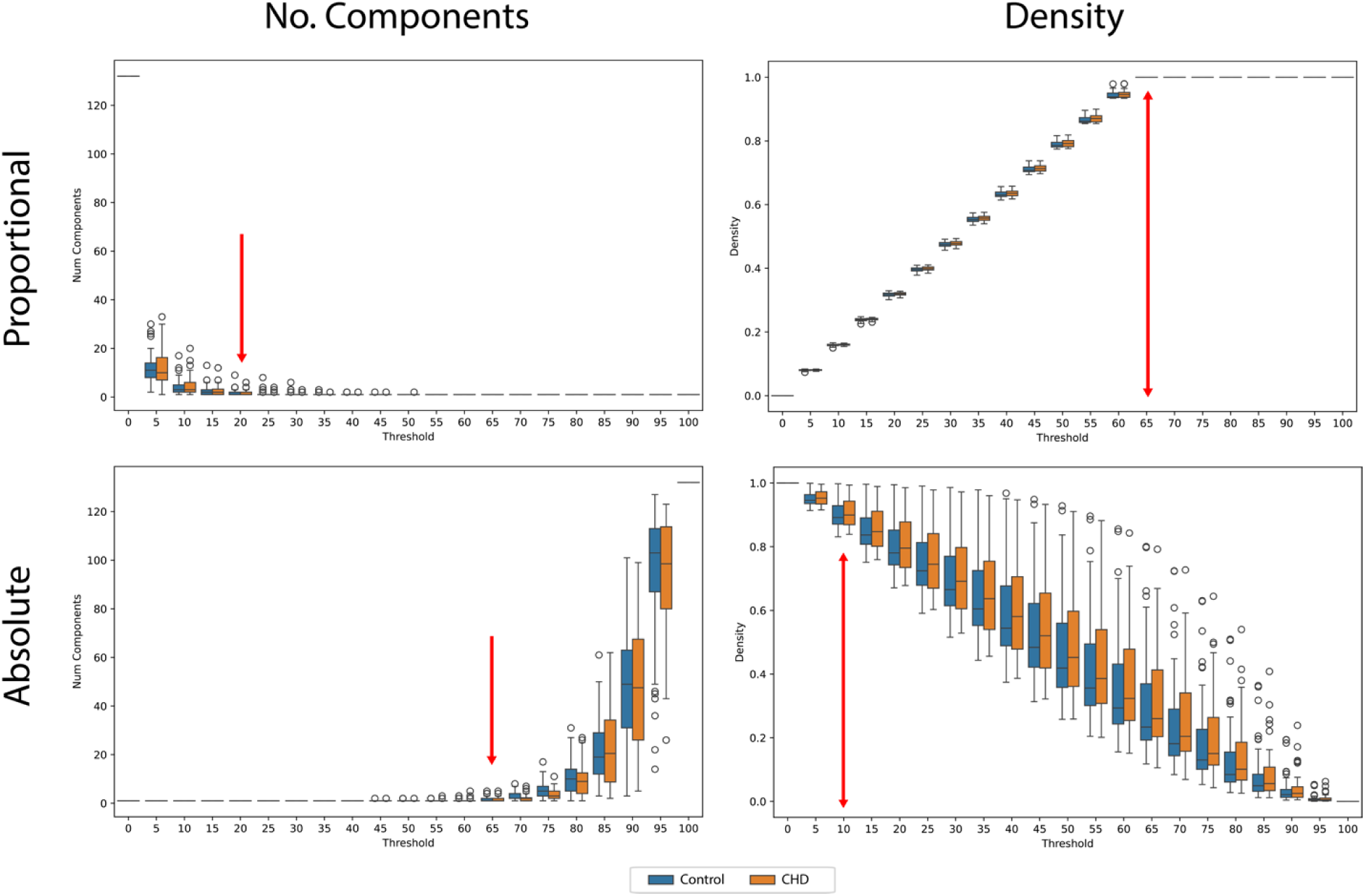
Absolute thresholding had a lower threshold of 0.10 to mitigate spurious connections and an upper threshold of 0.65 to limit the number of components in the network, an anatomical constraint. Proportional thresholding ranged from 0.20 to limit the number of components to 0.65, where all networks became fully connected. Note: A proportional threshold takes a top down approach where it selects the strongest X% of edges first, whereas an absolute thresholds take a bottom up approach keeping edges with strength >= X.

**Supplemental Figure 2.**
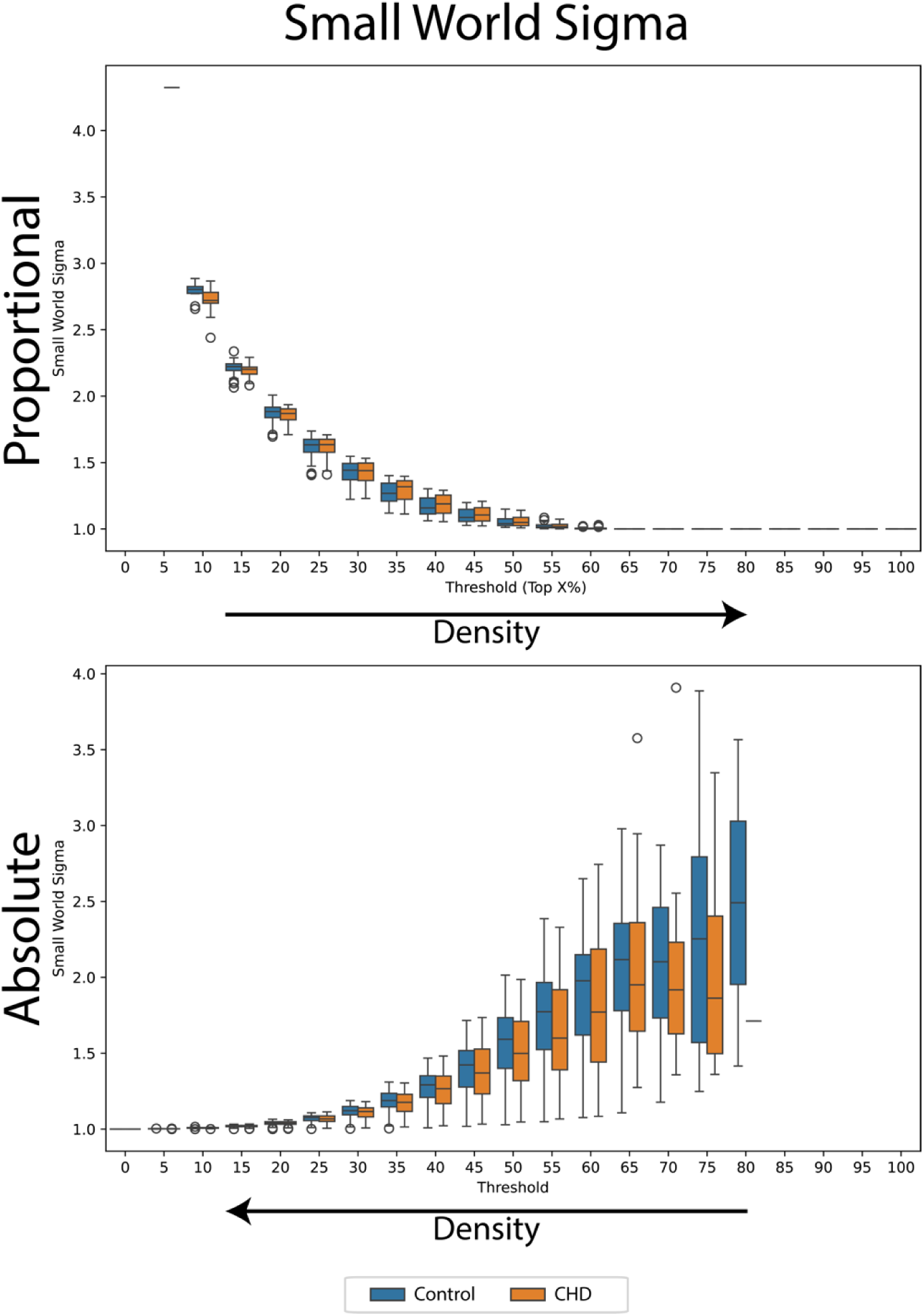
The small World Coefficient (sigma) tends to be larger in sparse graphs and decreases as thresholds become more lenient, resulting in denser networks. The Small World Coefficient was calculated only in networks with a single component. The notable difference in variance between absolute and proportional thresholding can be attributed to the fact that proportional thresholding controls for network density, whereas absolute thresholding does not, leading to highly variable edge counts in patient networks even at a single threshold.

